# Sociodemographic, health-related, and clinical characteristics and their associations with mortality among All of Us participants compared with the United States general population

**DOI:** 10.1101/2024.11.09.24317040

**Authors:** Jingxuan Wang, Erin L. Ferguson, Peter Buto, Ruijia Chen, Anna Pederson, Minhyuk Choi, Deborah Blacker, M. Maria Glymour

## Abstract

**Background:** The large and diverse All of Us Research Program offers tremendous opportunities for health research. However, results may not be generalizable to the US population due to the program’s targeted recruitment efforts.

**Methods:** We compared All of Us participants to those from the nationally representative 1999-2018 National Health and Nutrition Examination Survey (NHANES) with respect to: overall mortality rates; the distribution of sociodemographic, health-related, and clinical characteristics; the association of each characteristic with mortality estimated using Cox proportional-hazards models; and population attributable fractions (PAFs) for each characteristic and mortality.

**Results:** All of Us participants were older, less likely to be Non-Hispanic White, had more years of education, and had a higher prevalence of major chronic conditions than NHANES. Mortality rates were generally lower for All of Us participants, especially at older ages. The direction of associations in All of Us and NHANES matched for almost all comparisons, but differed in magnitude for some conditions, primarily clinical diagnoses. For example, in All of Us, mortality among participants with a prevalent cancer diagnosis was 2.79 (95% CI: 2.59 to 3.01) times higher than among participants without cancer; in NHANES the hazard ratio was only 1.24 (95% CI: 1.16 to 1.33). PAFs were generally higher in All of Us.

**Conclusions:** Predictors of mortality in All of Us do not consistently generalize to the US population. Analytical approaches are needed to address non-representativeness and mitigate potential biases associated with the selection into the All of Us cohort.

## Introduction

Large-scale volunteer databanks such as the All of Us Research Program collect extensive social, biological, and clinical data from hundreds of thousands of participants.[1–5] To achieve the desired sample size and diversity, databanks typically use a combination of targeted and convenience recruitment strategies. These *ad hoc* recruitment efforts stand in marked contrast to major health studies that recruit based on probability sampling from a defined population, such as the National Health and Nutrition Examination Survey, the National Health Interview Survey, or the Health and Retirement Study. Yet, the goal of biobank-based research is nearly always to generalize findings to a population. Although selection into a sample has the potential to undermine both external and internal validity, the statistical impact in practice sometimes turns out to be negligible.[6] Thus, it is essential to evaluate whether results in large studies such as All of Us mirror those in the overall population.

The All of Us Research Program exemplifies the challenges of such mega-studies. Launched in 2018, All of Us aimed to enroll at least 1 million U.S. adults, gather baseline data, and link to electronic health records (EHRs).[2] All of Us was born out of the National Institutes of Health Precision Medicine Initiative, and from its inception the principles of representative sampling were explicitly rejected.[7] Despite being relatively new, there are already 397 publications based on All of Us data (Figure 1). As with other large databanks, the advantages of All of Us with respect to large sample size and phenotype diversity came at the expense of non-representative enrollment and low response rates that further affect representativeness.[8,9]

**Figure 1.**
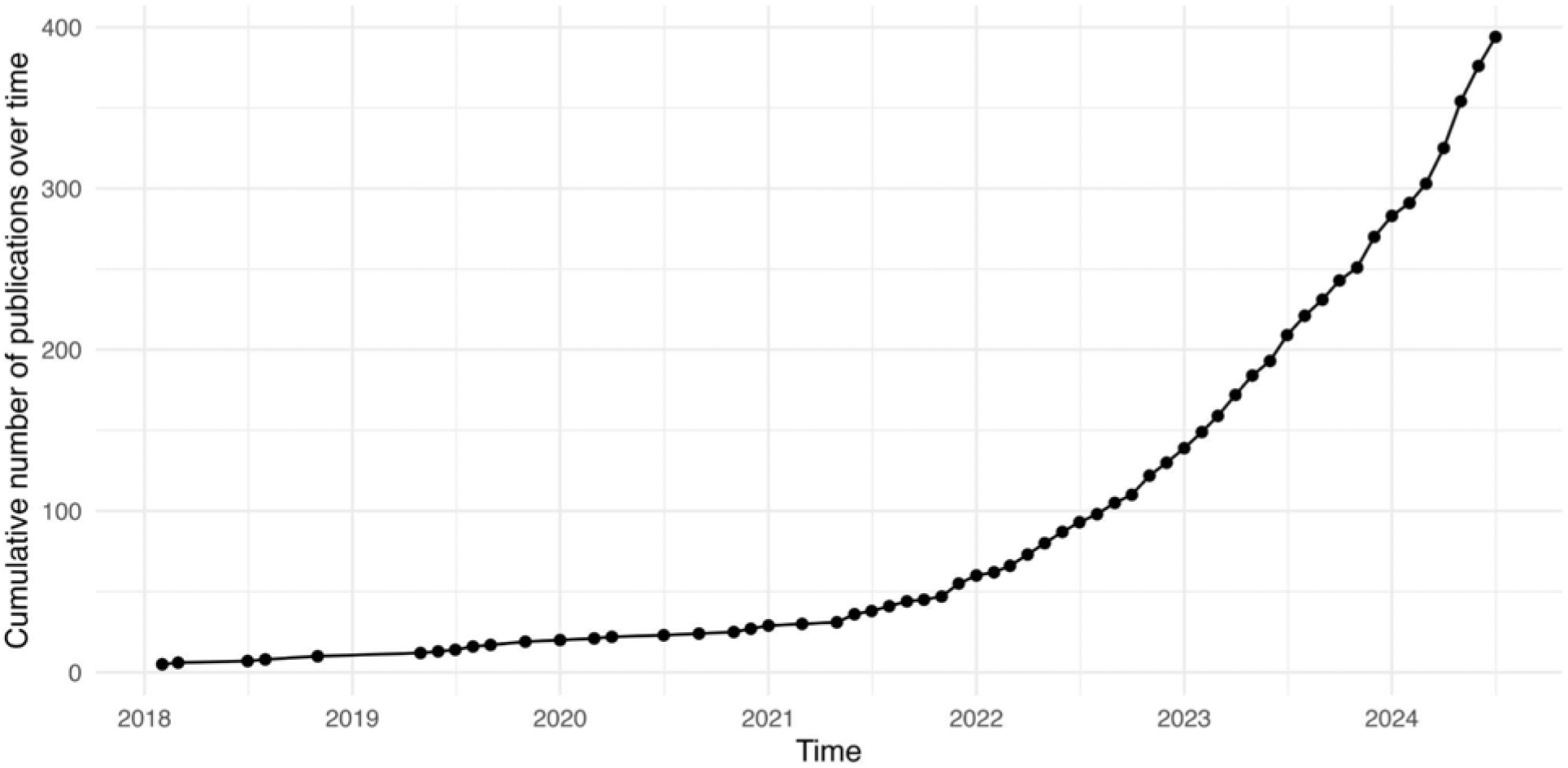
Number of publications over time based on All of Us.

To investigate whether evidence from All of Us can be generalized to U.S. adults, we compared the sociodemographic, health-related, and clinical characteristics of All of Us participants with those of the nationally representative continuous National Health and Nutrition Examination Survey (NHANES). We also evaluated the generalizability of risk factor-mortality associations on the hazard ratio scale and compared population attributable fractions (PAFs) for mortality estimated in the All of Us cohort to PAFs estimated in NHANES.

## Methods

### Study population and participants

The All of Us research program began recruiting individuals aged 18 and older living in the U.S. in 2017 through over 340 recruitment sites. Participants can directly sign up online, at recruitment events, or through one of the participating healthcare provider organizations (which include large academic medical centers, Veterans’ Administration medical centers, and community health centers). At enrollment, participants complete baseline surveys and have the option to take part in additional health surveys and ancillary studies, which may require in-person visits or biospecimen collection. They may also consent to the sharing of physical measurements and/or electronic health record (EHR) data. About 95% of participants have consented to share their EHR data to-date, enabling health outcomes to be followed longitudinally through linkage to their EHRs. All EHR data are harmonized across sites using the Observational Medical Outcomes Partnership (OMOP) Common Data Model.[10] However, not all participants who consented to share EHR have any EHR data available at the time of analysis. In the current manuscript, we restricted analyses to All of Us participants with EHR data available. (eFigure 1).

The continuous NHANES is a complex, multistage probability sample of the U.S. noninstitutionalized population that began in 1999, with a sampling methodology designed to ensure national representativeness. For this analysis, we included NHANES datasets from 1999 to 2018, which were collected across ten 2-year cycles (i.e., 1999-2000, 2001-2002, every 2 years through 2017-2018).[11] We restricted to participants over age 18 whose mortality status was available for public release.

### Assessment of baseline characteristics

We examined sociodemographic, health-related, and clinical indicators in both All of Us and continuous NHANES. All variables were recoded for consistency between the data sources and harmonization of measurements is detailed in eAppendix 1. Sociodemographic characteristics were self-reported in both samples: age, race/ethnicity (Non-Hispanic White, Non-Hispanic Black, Non-Hispanic Asian, Hispanic, other), sex (female, male, other), education (less than high school, high school, some college, college and above), household income (<10k, 10-25k, 25-35k, 35-75k, >75k), marital status (married, living with partner, divorced, separated, widowed, never married), and country of birth (U.S., outside U.S.). Health-related factors were also self-reported in both samples: smoking history (yes, no), alcohol history (yes, no), marijuana history (yes, no), serious hearing difficulty (yes, no), health insurance (yes, no), and self-assessed general health (excellent, very good, good, fair, poor).

Clinical indicators included diagnoses of asthma, coronary heart disease, congestive heart failure, heart attack, cancer, hypertension, obesity (body mass index [BMI] ≥ 30 kg/m^2^), stroke, diabetes mellitus, and cataract surgery. In All of Us, all clinical characteristics except for obesity were ascertained from EHRs based on diagnostic codes (eTable 1) recorded prior to the date of enrollment. In continuous NHANES, all clinical variables except for obesity were self-reported through questions “Has a doctor or other health professional ever told you that you had a certain condition.” Obesity was defined as a BMI greater than 30, with BMI calculated from measured height and weight.

### Mortality status identification

Death was documented in participants’ EHRs in All of Us. In continuous NHANES, mortality data was obtained through linkage with death certificate records from the National Death Index (NDI) up to December 31, 2019. In both datasets, we defined all-cause mortality as the primary outcome.

### Statistical analysis

We first compared the baseline characteristics of participants and age-specific mortality rates across All of Us and the continuous NHANES. We then estimated the associations between each characteristic and mortality using Cox proportional-hazard models, adjusting for baseline age, sex, and race/ethnicity when these variables were not the characteristic of interest. Participants were followed from enrollment to the date of death from any cause, the end of EHR follow-up in All of Us (eAppendix 3), or the administrative censoring for continuous NHANES (December 31, 2019), whichever came first. In continuous NHANES, we applied survey weights to all statistical analyses to generate estimates representative of the non-institutionalized civilian U.S. population. Given the large sample sizes, even small differences might be statistically significant. We selected a threshold of a 20% difference in the magnitude of the hazard ratios in All of Us versus NHANES as an important difference.

To further illustrate the importance of divergent results in the two studies for prioritizing public health targets, we then compared the corresponding PAF for each modifiable factor in All of Us and NHANES. Additionally, we calculated the PAF using the hazard ratio estimates from All of Us and the prevalence estimates from NHANES to better distinguish whether the observed differences in prioritization are driven by the hazard ratios, the prevalence, or both. PAFs were calculated using the Levin’s formula (formulas in eAppendix 3).[12,13]

Follow-up in EHR data may be incomplete, resulting in potential missed diagnoses in All of Us. We considered this unlikely for individuals with high health care utilization. We therefore repeated the associational analysis within All of Us in our secondary analysis, stratifying participants by low, middle, and high levels of healthcare utilization which was defined by tertiles of all EHR visits within one year before baseline. Healthcare utilization was defined based on the number of recorded encounters with the healthcare system in the EHR during the year prior to enrollment.

All statistical analyses were performed using R version 4.4.0.

## Results

### Baseline characteristics comparison

Among the 249,121 All of Us participants included in our analysis, the mean (SD) age was 53.3 years (16.9), while the weighted continuous NHANES participants averaged 46.3 years (0.1; Table 1). All of Us participants differed from the representative NHANES participants in nearly every characteristic assessed, although the differences varied across characteristics. For example, All of Us participants were less likely to have a history of smoking (38.3% vs. 44.7%), but more likely to have used alcohol (88.5% vs. 66.2%) or marijuana (48.6% vs. 27.5%). All of Us participants averaged higher education but were less likely to report “excellent” health (11.3% vs. 17.6%).

**Table 1.** Sociodemographic, health-related, and clinical characteristics of All of Us and continuous NHANES participants.

| Characteristic n (%)<sup>a</sup> | Continuous NHANES (unweighted) | Continuous NHANES (weighted) | All of Us (EHR sample) |
| --- | --- | --- | --- |
| N | 59,057 | 224,455,537 | 249,121 |
| ***Sociodemographic characteristics*** |  |  |  |
| Mean age (SD) | 48.0 (19.5) | 46.3 (0.1) | 53.3 (16.9) |
| Sex |  |  |  |
| Female | 30,574 (51.8%) | 116,309,932 (51.8%) | 152,210 (61.1%) |
| Male | 28,483 (48.2%) | 108,145,605 (48.2%) | 91,790 (36.8%) |
| Other or missing | 0 (0%) | 0 (0%) | 5,121 (2.1%) |
| Race/ethnicity |  |  |  |
| Non-Hispanic White | 25,378 (43.0%) | 151,683,379 (67.6%) | 141,007 (56.6%) |
| Non-Hispanic Asian | 3,080 (5.2%) | 5,259,512 (2.3%) | 8,055 (3.2%) |
| Non-Hispanic Black | 12,625 (21.4%) | 25,514,664 (11.4%) | 45,474 (18.3%) |
| Hispanic | 15,587 (26.4%) | 31,468,010 (14.0%) | 39,639 (15.9%) |
| Other | 2,387 (4.0%) | 10,529,973 (4.7%) | 8,007 (3.2%) |
| Missing | 0 (0%) | 0 (%) | 6,939 (2.8%) |
| Education |  |  |  |
| Less than high school | 16,829 (28.5%) | 40,723,708 (18.1%) | 19,882 (8.0%) |
| High school | 15,106 (25.6%) | 57,160,283 (25.5%) | 42,584 (17.1%) |
| Some college | 15,326 (26.0%) | 66,388,396 (29.6%) | 61,488 (24.7%) |
| College or above | 11,674 (19.8%) | 59,848,353 (26.7%) | 117,762 (47.3%) |
| Missing | 122 (0.2%) | 334,798 (0.1%) | 7,405 (3.0%) |
| Household income |  |  |  |
| Less than 10k | 4,092 (6.9%) | 10,323,215 (4.6%) | 30,115 (12.1%) |
| 10-25k | 12,797 (21.7%) | 35,625,516 (15.9%) | 27,780 (11.2%) |
| 25-35k | 6,773 (11.5%) | 22,006,422 (9.8%) | 17,094 (6.9%) |
| 35-75k | 16,037 (27.2%) | 66,561,558 (29.7%) | 46,552 (18.7%) |
| More than 75k | 12,730 (21.6%) | 69,478,209 (31.0%) | 79,399 (31.9%) |
| Missing | 6,628 (11.2%) | 20,460,618 (9.1%) | 48,181 (19.3%) |
| Marital status |  |  |  |
| Married | 28,454 (48.2%) | 119,299,931 (53.2%) | 111,263 (44.7%) |
| Living with partner | 4,134 (7.0%) | 16,225,518 (7.2%) | 14,833 (6.0%) |
| Divorced | 5,592 (9.5%) | 21,594,579 (9.6%) | 34,826 (14.0%) |
| Separated | 1,823 (3.1%) | 5,337,424 (2.4%) | 7,656 (3.1%) |
| Widowed | 5,038 (8.5%) | 13,339,218 (5.9%) | 14,326 (5.8%) |
| Never married | 11,601 (19.6%) | 41,485,073 (18.5%) | 58,099 (23.3%) |
| Missing | 2,415 (4.1%) | 7,173,793 (3.2%) | 8,118 (3.3%) |
| Country of birth |  |  |  |
| U.S. | 43,395 (73.5%) | 186,274,904 (83.0%) | 207,053 (83.1%) |
| Outside U.S. | 15,615 (26.4%) | 38,043,885 (16.9%) | 37,469 (15.0%) |
| Missing | 47 (0.1%) | 136,749 (0.1%) | 4,599 (1.8%) |
| **Health-related characteristics** |  |  |  |
| Smoking history |  |  |  |
| Yes | 25,091 (42.5%) | 100,389,865 (44.7%) | 95,504 (38.3%) |
| No | 30,682 (52.0%) | 118,440,003 (52.8%) | 146,565 (58.8%) |
| Missing | 3,284 (2.5%) | 5,625,669 (2.5%) | 7,052 (2.8%) |
| Alcohol history |  |  |  |
| Yes | 34,750 (58.8%) | 148671716 (66.2%) | 220,548 (88.5%) |
| No | 13,887 (23.5%) | 44966008 (20.0%) | 23,565 (9.5%) |
| Missing | 10,420 (17.6%) | 30817813 (13.7%) | 5,008 (2.0%) |
| Marijuana history |  |  |  |
| Yes | 12,578 (21.3%) | 61,802,725 (27.5%) | 121,170 (48.6%) |
| No | 10,936 (18.5%) | 42,013,904 (18.7%) | 112,904 (45.3%) |
| Missing | 35,543 (60.2%) | 120,638,909 (53.7%) | 15,047 (6.0%) |
| Serious hearing difficulty |  |  |  |
| Yes | 3,119 (5.3%) | 10,109,144 (4.5%) | 6,081 (2.4%) |
| No | 55,911 (94.7%) | 214,283,439 (95.5%) | 80,242 (32.2%) |
| Missing | 27 (0.0%) | 62,954 (0.0%) | 162,798 (65.3%) |
| Health insurance |  |  |  |
| Yes | 46,210 (78.2%) | 183,604,536 (81.8%) | 229,687 (92.2%) |
| No | 12,459 (21.1%) | 39,636,682 (17.7%) | 11,867 (4.8%) |
| Missing | 388 (0.7%) | 1,214,319 (0.5%) | 7,567 (3.0%) |
| General health |  |  |  |
| Excellent | 8,953 (15.2%) | 39,426,851 (17.6%) | 28,225 (11.3%) |
| Very good | 15,466 (26.2%) | 70,132,205 (31.2%) | 77,802 (31.2%) |
| Good | 20,687 (35.0%) | 75,275,841 (33.5%) | 82,382 (33.1%) |
| Fair | 11,164 (18.9%) | 31,959,853 (14.2%) | 45,845 (18.4%) |
| Poor | 2,740 (4.6%) | 7,482,779 (3.3%) | 11,282 (4.5%) |
| Missing | 47 (0.1%) | 178,009 (0.1%) | 3,585 (1.4%) |
| ***Clinical characteristics*** |  |  |  |
| Asthma |  |  |  |
| Yes | 7,985 (13.5%) | 31,618,163 (14.1%) | 35,726 (14.3%) |
| No | 51,014 (86.4%) | 192,620,910 (85.8%) | 213,395 (85.7%) |
| Missing | 58 (0.1%) | 216,465 (0.1%) | 0 (0%) |
| Coronary heart disease |  |  |  |
| Yes | 2,345 (4.0%) | 7,506,504 (3.3%) | 23,415 (9.4%) |
| No | 52,329 (88.6%) | 208,560,907 (92.9%) | 225,706 (90.6%) |
| Missing | 4,383 (7.4%) | 8,388,126 (3.7%) | 0 (0%) |
| Congestive heart failure |  |  |  |
| Yes | 1,903 (3.2%) | 5,250,324 (2.3%) | 10,775 (4.3%) |
| No | 52,854 (89.5%) | 211,098,929 (94.0%) | 238,346 (95.7%) |
| Missing | 4,300 (7.3%) | 8,106,284 (3.6%) | 0 (0%) |
| Heart attack |  |  |  |
| Yes | 2,458 (7.2%) | 7,433,340 (3.3%) | 9,716 (3.9%) |
| No | 52,376 (88.7%) | 209,045,498 (93.1%) | 239,405 (96.1%) |
| Missing | 4,223 (4.2%) | 7,976,699 (3.6%) | 0 (0%) |
| Cancer |  |  |  |
| Yes | 5,161 (8.7%) | 20,686,450 (9.2%) | 39,098 (15.7%) |
| No | 49,718 (84.2%) | 195,832,202 (87.2%) | 210,023 (84.3%) |
| Missing | 4,178 (7.1%) | 7,936,885 (3.5%) | 0 (0%) |
| Hypertension |  |  |  |
| Yes | 19,265 (32.6%) | 66,042,784 (29.4%) | 92,276 (37.0%) |
| No | 39,487 (66.9%) | 157,641,644 (70.2%) | 156,845 (63.0%) |
| Missing | 305 (0.5%) | 771,110 (0.3%) | 0 (0%) |
| Obesity |  |  |  |
| Yes | 19,306 (32.7%) | 73,374,367 (32.7%) | 101,152 (40.6%) |
| No | 35,792 (60.6%) | 137,870,972 (61.4%) | 147,969 (59.4%) |
| Missing | 3,959 (6.7%) | 13,210,199 (5.9%) | 0 (0%) |
| Stroke |  |  |  |
| Yes | 2,191 (3.7%) | 6,148,920 (2.7%) | 602 (0.2%) |
| No | 52,667 (89.2%) | 210,352,885 (93.7%) | 248,519 (99.8%) |
| Missing | 4,199 (7.1%) | 7,953,733 (3.5%) | 0 (0%) |
| Diabetes mellitus |  |  |  |
| Yes | 6,688 (11.3%) | 19,148,057 (8.5%) | 41,814 (16.8%) |
| No | 51,219 (86.7%) | 201,223,879 (89.6%) | 207,307 (83.2%) |
| Missing | 1,150 (1.9%) | 4,083,601 (1.8%) | 0 (0%) |
| Cataract surgery |  |  |  |
| Yes | 2,024 (3.4%) | 5,162,358 (2.3%) | 970 (0.4%) |
| No | 23,382 (39.6%) | 90,277,751 (40.2%) | 248,151 (99.6%) |
| Missing | 33,651 (57.0%) | 129,015,429 (57.5%) | 0 (0%) |

Abbreviations: SD, standard deviation.
<sup>a</sup> Percentage may not sum to 100 due to rounding.

### Mortality rate comparison

In All of Us, 3,216 deaths were recorded over a median follow-up of 2.8 years. In the continuous NHANES, 9,242 deaths were recorded over a median follow-up of 9.4 years. After applying weighting to the continuous NHANES, we observed 25,665,442 deaths with a median follow-up of 9.6 years. The age-specific mortality rate in All of Us was slightly higher for the 40-50 age group, while it was lower for all other age groups compared to NHANES, with the differences becoming more pronounced at older ages (Figure 2, eTable 3). For example, the mortality rates for participants aged 50-60 were 4.4 per 1,000 person-years in All of Us, compared to 5.5 in NHANES. For those aged 60-70, the rates were 6.1 in All of Us and 11.7 in NHANES. Among participants aged 80 and above, the difference increased significantly, with rates of 17.5 in All of Us and 93.0 in NHANES, respectively.

**Figure 2.**
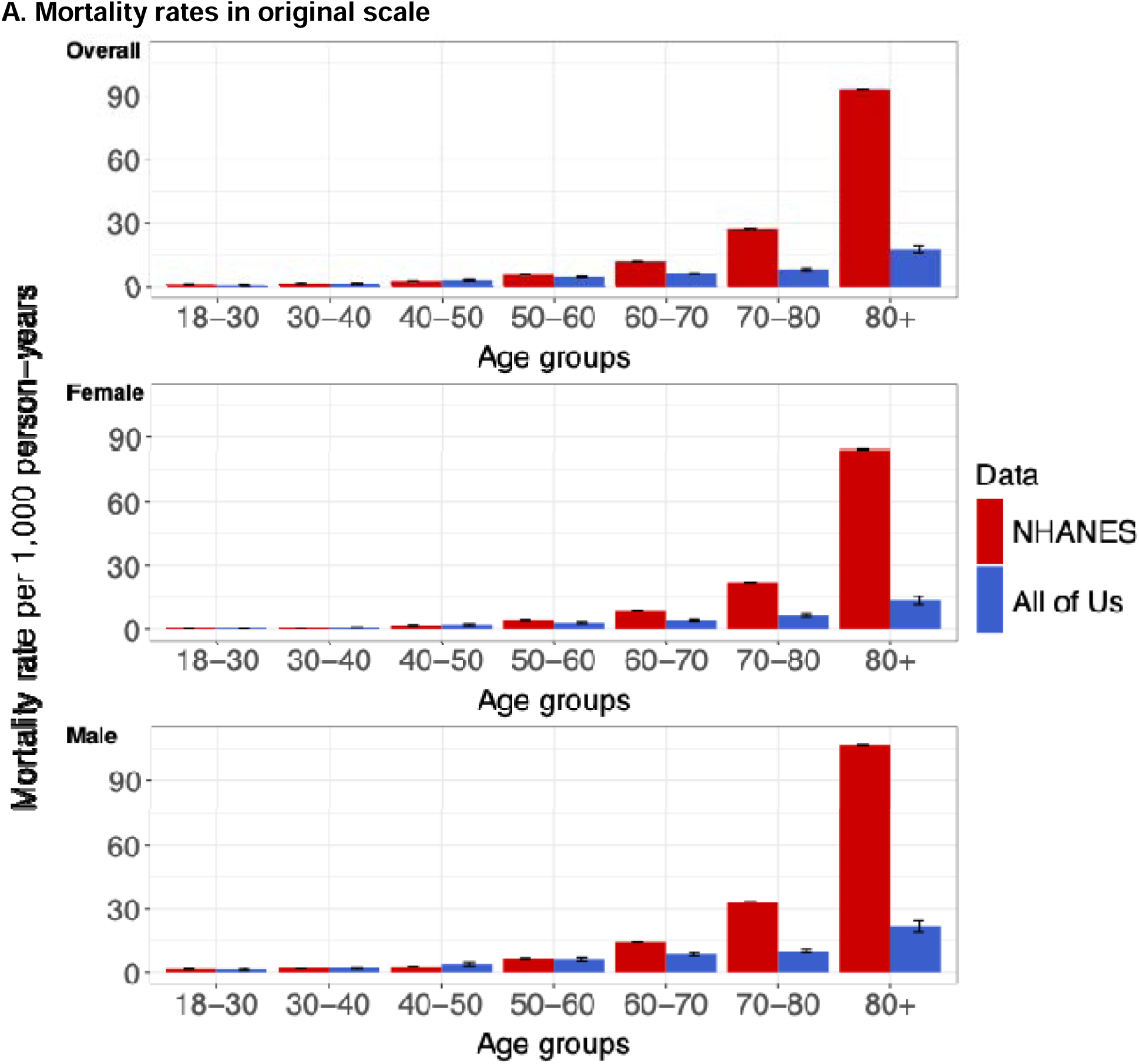
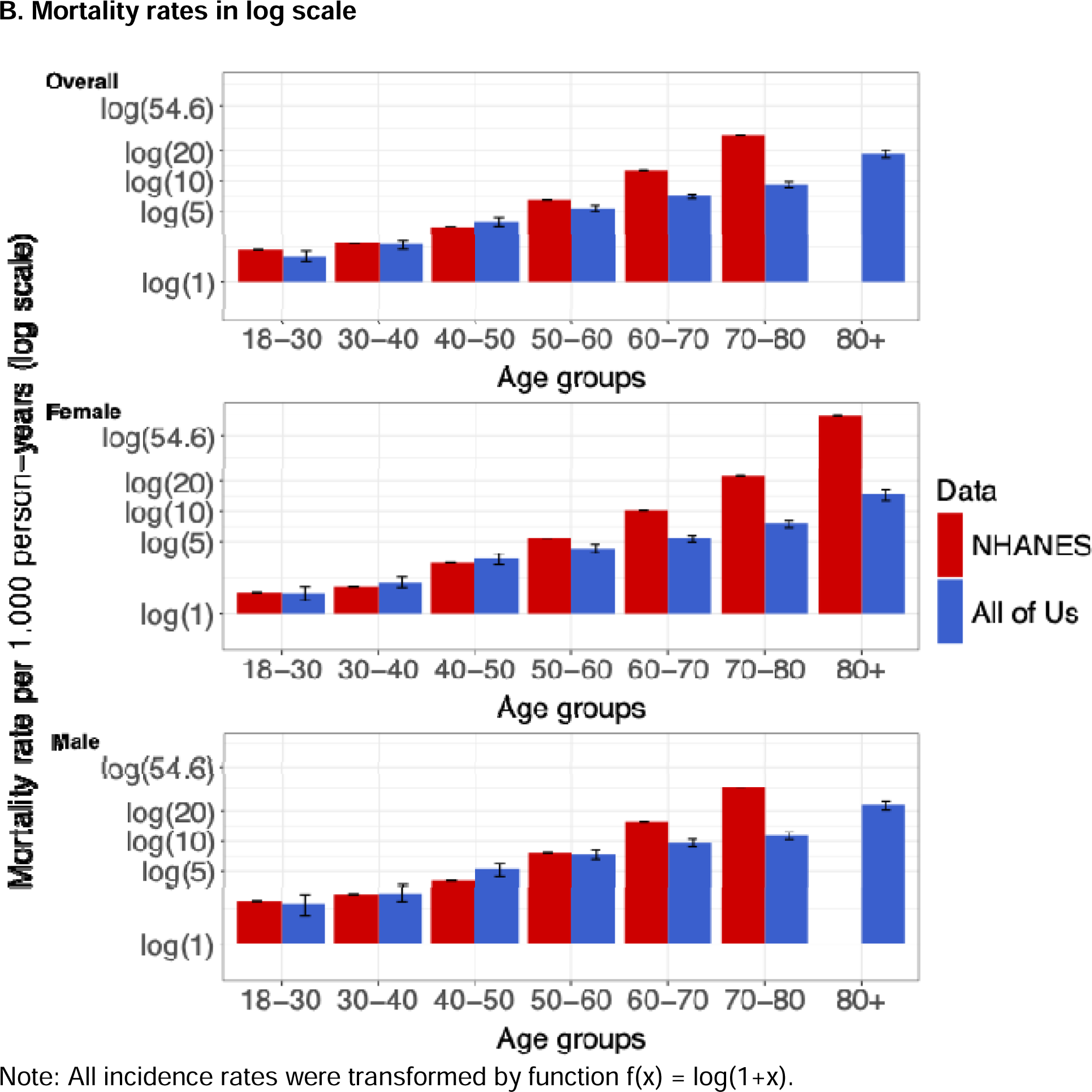
Age-specific mortality rates by sex in (A) original scale and (B) log scale in All of Us and continuous NHANES.

### Sociodemographic factors and mortality rate comparison

Across 23 comparisons made for the 7 sociodemographic factors, the magnitude of the hazard ratio differed by more than 20% for 11 comparisons and was in the opposite direction for 4 comparisons (Figure 3A). Age was a weaker risk factor for mortality in All of Us compared to the general population. In All of Us, the hazard ratio per decade increase in age was 1.59 (95% CI: 1.55 to 1.64), compared to 2.50 (95% CI: 2.44 to 2.56) in NHANES. The mortality rate for Non-Hispanic Asian participants was similar to that for Non-Hispanic White participants in All of Us (HR=0.95, 95% CI: 0.73 to 1.24), while the mortality rate was lower for Non-Hispanic Asian participants in NHANES (HR=0.53, 95% CI: 0.41 to 0.67). Hispanic participants had a higher mortality hazard compared to Non-Hispanic White participants in All of Us (HR=1.24, 95% CI: 1.11 to 1.39), while the mortality hazard was lower for Hispanic participants in NHANES participants (HR=0.87, 95% CI: 0.80 to 0.95). The hazard ratio for being male was larger in All of Us (HR=1.78, 95% CI: 1.66 to 1.91) compared to NHANES participants (HR=1.45, 95% CI: 1.37 to 1.52). The associations between education and mortality were similar in the two datasets, except that in All of Us, the mortality rate for participants who completed high school did not differ from those who did not (HR=0.91, 95% CI: 0.80 to 1.03). The magnitudes of associations between income and mortality were similar in All of Us and NHANES. For example, the mortality hazard for participants with an annual household income more than $75,000 was 0.68 times the hazard for those with $35,000-$75,000 income in All of Us (95% CI: 0.60 to 0.76) and 0.65 (95% CI: 0.59 to 0.72) in NHANES. In All of Us, living with a partner and never being married were associated with lower hazard ratios compared to being married, while the hazard ratios for being separated, divorced, or widowed were similar to those in NHANES. The hazard ratio for being born outside the U.S. was higher in All of Us (HR=0.83, 95% CI: 0.73 to 0.95) than NHANES (HR=0.64, 95% CI: 0.69 to 0.71).

**Figure 3.**
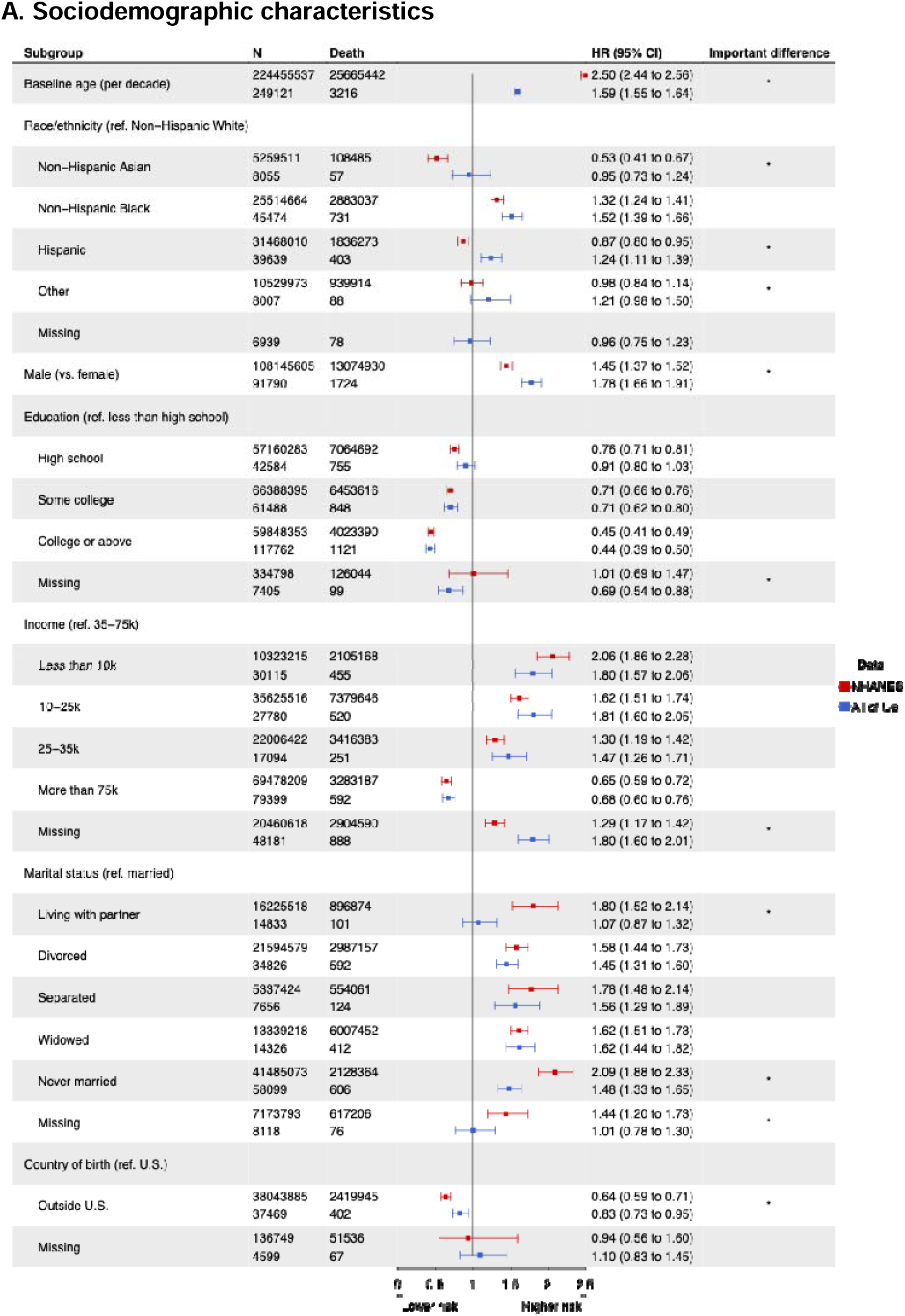
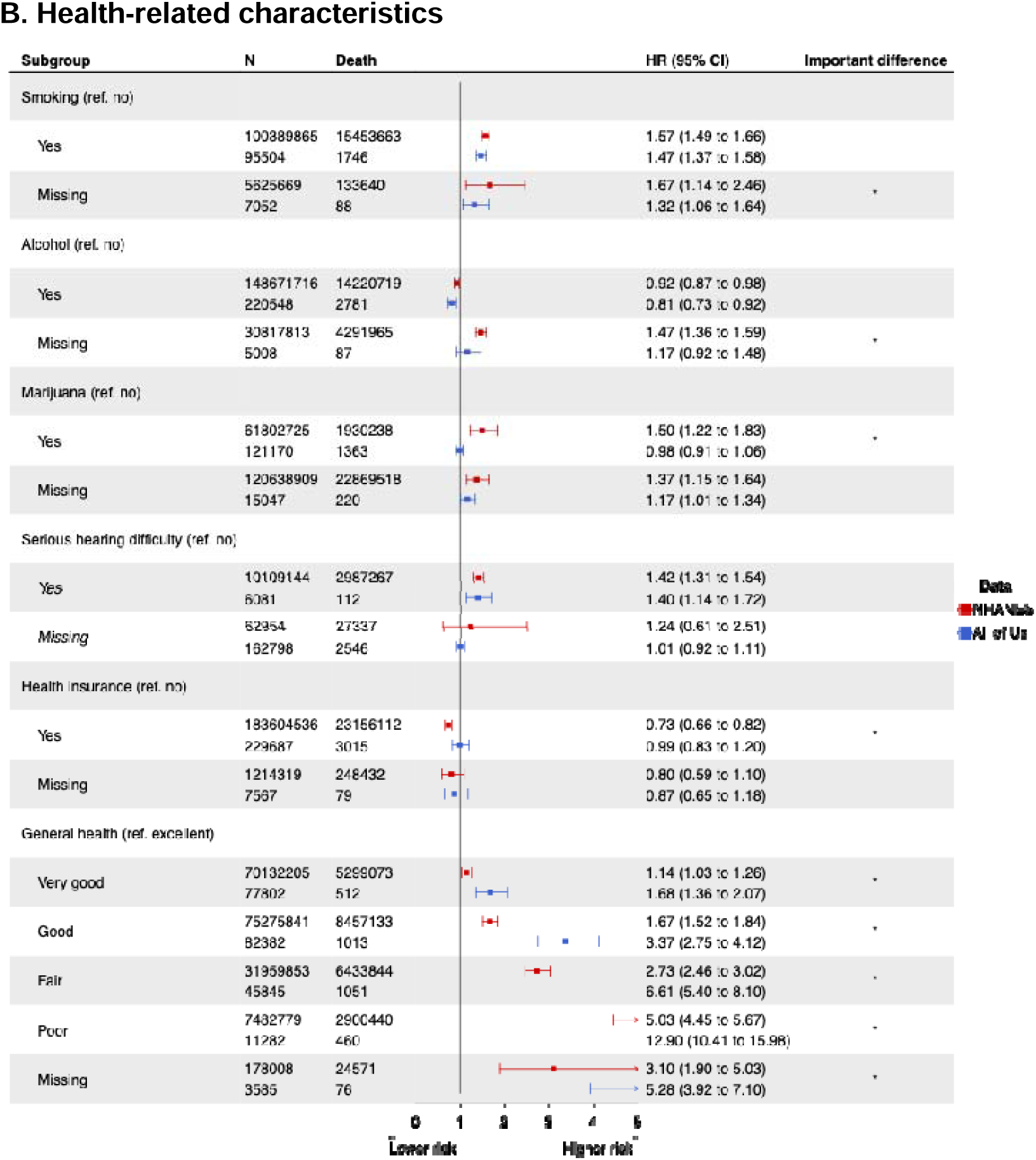
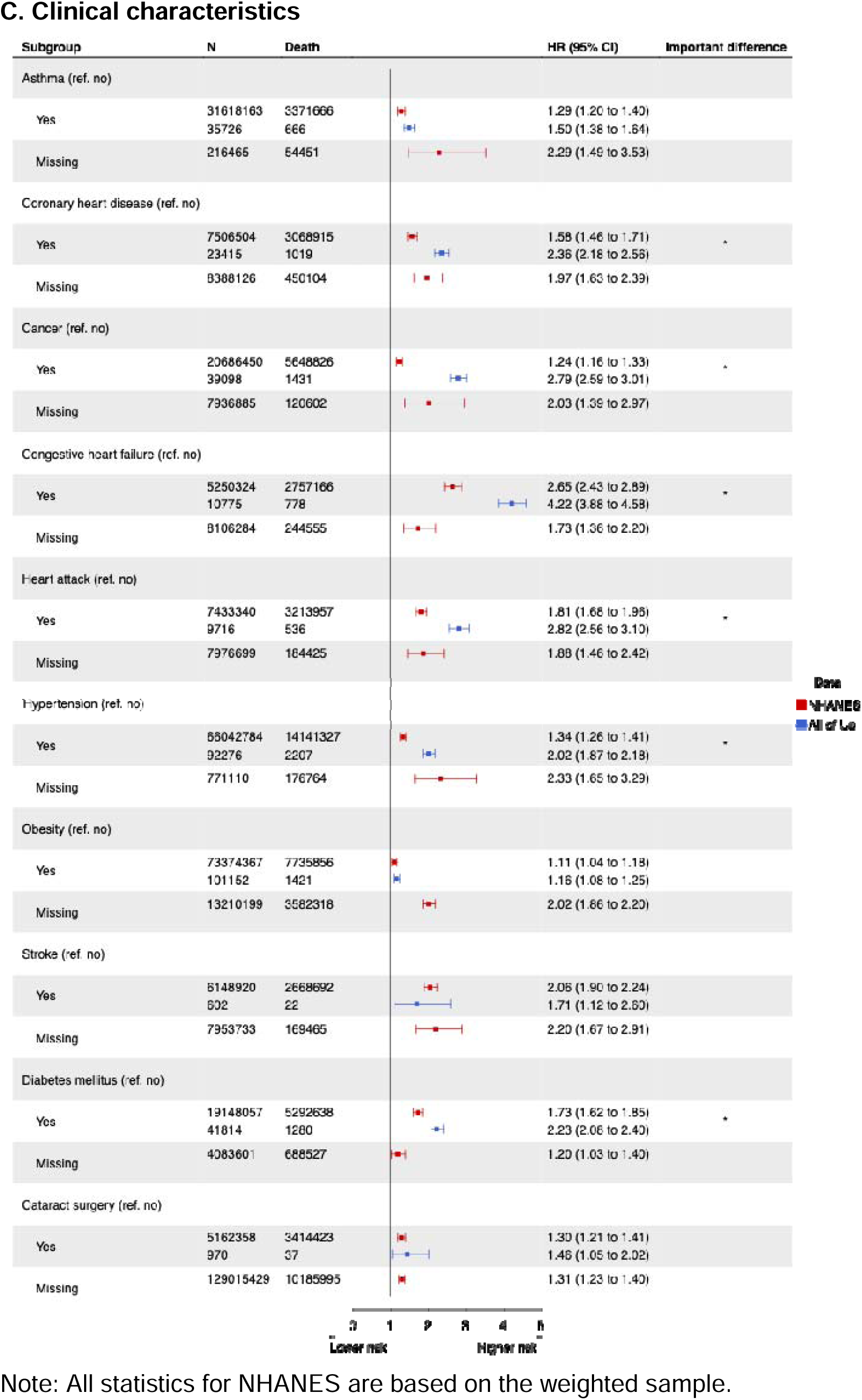
Associations of baseline (A) sociodemographic, (B) health-related, and (C) clinical characteristics with mortality in All of Us and continuous NHANES.

### Health-related factors and mortality rate comparison

Across 14 comparisons for 6 health-related risk factors, 9 differed by more than 20% between All of Us and NHANES, and 1 had a qualitatively opposite estimate (Figure 3B). The hazard ratios for smoking, alcohol, and serious hearing difficulty did not significantly differ in All of Us and NHANES. Marijuana use history was not associated with mortality in All of Us (HR=0.98, 95% CI: 0.91 to 1.06), but it was linked to higher mortality in NHANES (HR=1.50, 95% CI: 1.22 to 1.83). Having health insurance was only associated with a lower mortality in NHANES (HR=0.99, 95% CI: 0.83 to 1.20 in All of Us; HR=0.73, 95% CI: 0.66 to 0.82 in NHANES). The associations between self-reported general health and mortality were stronger in All of Us than in NHANES. For example, the hazard ratio comparing participants reporting poor to those reporting excellent health was 12.90 (95% CI: 10.41 to 15.98) in All of Us, compared to 5.03 (95% CI: 4.45 to 5.67) in NHANES.

### Clinical factors and mortality rate comparison

Across 10 comparisons for 10 clinical risk factors, 6 differed by more than 20% between All of Us and NHANES, but none had qualitatively opposite estimates (Figure 3C). Asthma, obesity, stroke, and cataract surgery had similar hazard ratios for mortality in the two datasets, while the associations of coronary heart disease, congestive heart failure, cancer, heart attack, hypertension, and diabetes mellitus with mortality were stronger in All of Us. For example, in All of Us, cancer diagnosis was associated with 2.79 times higher hazard (95% CI: 2.59 to 3.01) for mortality, while the hazard ratio was 1.24 (95% CI: 1.16 to 1.33) in NHANES.

Stratifying by the low, middle, and high levels of healthcare utilization substantially changed the results in All of Us (eFigure 3). In general, the associations between characteristics and mortality in All of Us were weaker among participants with higher healthcare utilization compared to those with lower utilization. For example, the hazard ratio for diabetes mellitus decreased from 3.02 (95% CI: 2.26 to 4.03) in participants with low healthcare utilization to 1.75 (95% CI: 1.61 to 1.91) in those with high utilization in All of Us.

### Population attributable fractions comparison

PAFs for mortality due to sociodemographic factors were either similar or lower in All of Us (Table 2). For example, the PAF for low education was 31.0% in All of Us and 36.3% in NHANES. We observed divergent public health prioritization based on PAF regarding health-related characteristics. PAFs for smoking (15.7% vs. 20.8%) and no health insurance (0.0% vs. 6.0%) were lower in All of Us than in NHANES, while those for no alcohol use (2.1% vs. 1.9%), serious hearing difficulty (2.7% vs. 1.9%), and worse general health (72.3% vs. 39.4%) were higher than NHANES. The difference between PAFs estimated in two datasets varied across clinical conditions, with especially large divergence for cancer (22.0% vs. 2.3%) and diabetes mellitus (17.2% vs. 6.0%). PAFs using prevalence estimates for NHANES and hazard ratio estimates from All of Us were generally more similar to PAFs in NHANES, compared to PAFs in All of Us but still diverged (eTable4).

**Table 2.** Population attributable fraction for mortality due to characteristics estimated in All of Us and continuous NHANES.

| Characteristic<br>(reference category) | Continuous NHANES | All of Us |
| --- | --- | --- |
| ***Sociodemographic characteristics*** |  |  |
| Education (College or above) | 36.3% | 31.0% |
| Household income (More than 75k) | 39.3% | 40.7% |
| Marital status (Married) | 27.7% | 19.4% |
| Country of birth (Outside U.S.) | 31.4% | 14.6% |
| ***Health-related characteristics*** |  |  |
| Smoking history (No) | 20.8% | 15.7% |
| Alcohol history (Yes) | 1.9% | 2.1% |
| Serious hearing difficulty (No) | 1.9% | 2.7% |
| Health insurance (Yes) | 6.0% | 0.0% |
| General health (Excellent) | 39.4% | 72.3% |
| ***Clinical characteristics*** |  |  |
| Asthma (No) | 4.0% | 6.7% |
| Coronary heart disease (No) | 2.0% | 11.3% |
| Congestive heart failure (No) | 3.9% | 12.2% |
| Heart attack (No) | 2.7% | 6.6% |
| Cancer (No) | 2.3% | 22.0% |
| Hypertension (No) | 9.0% | 27.4% |
| Obesity (No) | 3.7% | 6.2% |
| Stroke (No) | 2.9% | 0.2% |
| Diabetes mellitus (No) | 6.0% | 17.2% |
| Cataract surgery (No) | 1.6% | 0.2% |

## Discussion

The characteristics of All of Us participants diverged substantially from the U.S. general population, often in ways that were not easy to anticipate. Mortality rates were lower for All of Us participants especially over age 60.[14,15] Many characteristics showed similar associations with all-cause mortality in the All of Us study compared to NHANES and the sign of associations matched for almost all comparisons. The magnitude of associations diverged for other factors however, particularly regarding clinical conditions. Consequently, the PAFs for mortality due to these clinical conditions in the All of Us are generally higher than in the general population.

The composition of the All of Us research program may be strongly influenced by the recruitment of participants through health provider organizations. Recruitment through clinical care systems might naturally lead to a higher representation of individuals with major clinical conditions in the cohort.[10] Our findings are consistent with prior evidence suggesting that despite the non-representativeness of volunteer-based cohort studies, these cohorts can be useful for health research, but are not consistently generalizable.[1,8,16–21] For example, UK Biobank participants exhibited lower mortality and cancer incidence over follow-up, suggesting a potential healthy volunteer selection bias. In contrast, All of Us participants are more likely to have clinical conditions such as coronary heart disease, heart failure, cancer, hypertension, obesity, and diabetes. The divergence from the UK Biobank patterns likely reflects the All of Us recruitment strategies centered on reaching participants via health provider organization, and thus oversampled participants with higher disease burdens and greater exposure to negative social determinants of health.[10,22] Despite higher prevalence of several health conditions, mortality rates for older adults in All of Us are lower than in the general population. This may suggest potential measurement errors due to missing death records in EHRs.

The relative associations of several clinical conditions with mortality were stronger in All of Us compared to the general population. Although the exposures we defined were prevalent conditions in both All of Us and NHANES, the All of Us recruitment at academic medical centers may lead to an overrepresentation of individuals *currently receiving care* for the condition. Individuals who are currently receiving care for a chronic condition—and at an academic medical center may be at higher mortality risk than individuals in the general population with a history of that condition.

A major concern in All of Us—which we could not directly evaluate—is the possibility of a dual selection process differentially enrolling the most ill among individuals with a chronic condition (due to recruitment via academic medical centers and health care providers) but the healthiest among individuals without a chronic condition (the typical healthy volunteer effect). Although this cannot be directly assessed, the stronger association between self-reported general health and mortality in All of Us compared to NHANES highlights this issue. This work also highlights potential challenges in the All of Us data, including incomplete EHR coverage and possibly missing death records.

The higher prevalence of clinical conditions in All of Us, combined with their stronger associations with mortality, explains the elevated PAFs for mortality due to these conditions in All of Us. This finding indicates that PAF estimates can substantially misrepresent public health priorities when based on convenience sampling and recruitment strategies. The PAFs were somewhat improved when using prevalence estimates from representative data. This is a critical caveat when interpreting results from the cohort: they should not be used directly to guide public health priorities based on population impact.

Our study has several limitations. First, the death records data were obtained from participants’ EHRs in All of Us and there might be missing death records if participants’ deaths were not recognized in the EHRs, leading to underestimates in mortality rates. A similar limitation applies to the identification of clinical conditions, where low healthcare utilization or seeking care outside of organizations with EHR releases could lead to an underestimation of prevalence. Second, the follow-up period in All of Us is shorter compared to NHANES, leading to loss of precision and potential period effects. Third, we did not include every clinical condition, so our conclusions may not extend to other characteristics. For example, the prevalence of several psychiatric disorders is lower than in the general population.[23] Fourth, imperfect harmonization of complex variables, particularly those derived from different sources—such as questionnaires versus EHRs—may account for some of the observed differences and contribute to larger discrepancies between two datasets. Fifth, we lacked longitudinal follow-up for NHANES participants, except for mortality data, which prevents us from investigating the incidence of clinical conditions. This limits our ability to compare the characteristics of the general population across multiple dimensions between the two datasets. Lastly, while the continuous NHANES surveys are designed to be nationally representative, survey respondents may nonetheless be healthier than the general population.[24]

In conclusion, our results suggest caution when interpreting results from All of Us. Sociodemographic, health-related, and clinical characteristics in All of Us, along with their associations with mortality and PAFs, are not consistently generalizable to the U.S. population. It is likely All of Us oversampled participants with certain clinical conditions, and the healthy counterparts in the cohort may be healthier than those in the general population. While large samples in databanks such as All of Us can produce statistically significant results with precise confidence intervals, selection bias may prevent generalizing to other populations. Caution is warranted when interpreting and generalizing results from such datasets. Future studies should consider analytical approaches to address non-representativeness and mitigate the biases that arise from it.[25–29]

## Supporting information

eAppendix

## Data Availability

All data produced are available online at https://allofus.nih.gov/ and https://www.cdc.gov/nchs/nhanes/index.htm

