## Supplementary material for "Sociodemographic, health-related, and clinical characteristics and their associations with mortality among All of Us participants compared with the United States general population": eAppendix

**eAppendix 1. Harmonization of characteristics between All of Us and continuous NHANES**

In this appendix, we provide detailed information on item wording and coding in All of Us and NHANES.

| Characteristic | All of Us | Continuous NHANES | Harmonized data |
| --- | --- | --- | --- |
| ***Sociodemographic characteristics*** | | | |
| Age | continuous age, calculated by subtracting date of birth from date of enrollment. Date of birth was generalized to the year of birth, with all participants assigned a standardized birth date of June 15. | continuous age, top coded at 85 (1999-2000 to 2005-2006) or 80 (2007-2008 to 2017-2018) | continuous age |
| Sex | *What was your biological sex assigned at birth?*  ● Female  ● Male  ● Intersex  ● None of these  ● Prefer not to answer | ● Female  ● Male | ● Female  ● Male  ● Other or missing |
| Race/ethnicity | Race:  ● Asian  ● Black or African American  ● Middle Eastern or North African  ● More than one population  ● Native Hawaiian or Other Pacific Islander  ● None of these  ● White  ● Prefer not to answer  Ethnicity:  ● Hispanic or Latino  ● Not Hispanic or Latino  ● Prefer not to answer | ● Mexican American  ● Other Hispanic  ● Non-Hispanic White  ● Non-Hispanic Black  ● Non-Hispanic Asian  ● Other | ● Non-Hispanic White  ● Non-Hispanic Asian  ● Non-Hispanic Black  ● Hispanic  ● Other |
| Education | *What is the highest grade or year of school you completed?*  ● Advanced Degree (=College or above in harmonized variable)  ● College Graduate (=College or above)  ● College One to Three (=Some college)  ● Twelve or GED (=High school)  ● Nine Through Eleven (=Less than high school)  ● Five Through Eight (=Less than high school)  ● One Through Four (=Less than high school)  ● Never Attended (=Less than high school)  ● Prefer not to answer | ● Less Than 9th Grade (=Less than high school in harmonized variable)  ● 9-11th Grade (Includes 12th grade with no diploma) (=Less than high school)  ● High School Grad/GED or Equivalent (=High school)  ● Some College or AA degree (=Some college)  ● College Graduate or above (=College or above) | ● Less than high school  ● High school  ● Some college  ● College or above |
| Household income | ● Less than 10k  ● 10k-25k  ● 25k-35k  ● 35k-50k  ● 50k-75k  ● 75k-100k  ● 100k-150k  ● 150k-200k  ● More than 200k  ● Prefer not to answer | ● $ 0 to $ 4,999  ● $ 5,000 to $ 9,999  ● $10,000 to $14,999  ● $15,000 to $19,999  ● $20,000 to $24,999  ● $25,000 to $34,999  ● $35,000 to $44,999  ● $45,000 to $54,999  ● $55,000 to $64,999  ● $65,000 to $74,999  ● $75,000 and Over  ● $75,000 to $99,999  ● $100,000 and Over | ● Less than 10k  ● 10-25k  ● 25-35k  ● 35-75k  ● More than 75k |
| Marital status | ● Married  ● Living with partner  ● Divorced  ● Separated  ● Widowed  ● Never married  ● Prefer not to answer | ● Married  ● Living with partner  ● Divorced  ● Separated  ● Widowed  ● Never married | ● Married  ● Living with partner  ● Divorced  ● Separated  ● Widowed  ● Never married |
| Country of birth | ● USA  ● Other  ● Prefer not to answer | ● Born in 50 US States or Washington, DC (=U.S. in harmonized variable)  ● Born in Mexico (=Outside U.S.)  ● Born in Other Spanish Speaking Country (=Outside U.S.)  ● Born in Other Non-Spanish Speaking Country (=Outside U.S.)  ● Others | ● U.S.  ● Outside U.S. |
| ***Health-related characteristics*** | | | |
| Smoking history | *Have you smoked at least 100 cigarettes in your entire life?*  ● Yes  ● No  ● Prefer not to answer  ● Don't know | *{Have you/Has SP} smoked at least 100 cigarettes in {your/his/her} entire life?*  ● Yes  ● No  ● Refused  ● Don't know | ● Yes  ● No |
| Alcohol history | *In your entire life, have you had at least 1 drink of any kind of alcohol, not counting small tastes or sips? (By a “drink,” we mean a can or bottle of beer, a glass of wine or a wine cooler, a shot of liquor, or a mixed drink with liquor in it.)*  ● Yes  ● No  ● Prefer not to answer | *Included are liquor (such as whiskey or gin), beer, wine, wine coolers, and any other type of alcoholic beverage. In any one year, {have you/has SP} had at least 12 drinks of any type of alcoholic beverage? By a drink, I mean a 12 oz. beer, a 4 oz. glass of wine, or an ounce of liquor.*  ● Yes  ● No  ● Refused  ● Don't know | ● Yes  ● No |
| Marijuana history | *In your lifetime, which of the following substances have you ever used?*   - *Marijuana (cannabis, pot, grass, hash, weed, etc.)*   ● Yes  ● No  ● Prefer not to answer | *Marijuana is also called pot or grass. Marijuana is usually smoked, either in cigarettes, called joints, or in a pipe. It is sometimes cooked in food. Hashish is a form of marijuana that is also called 'hash.' It is usually smoked in a pipe. Another form of hashish is hash oil. Have you ever, even once, used marijuana or hashish? (available since 2005-2006)*  ● Yes  ● No  ● Refused  ● Don't know | ● Yes  ● No |
| Serious hearing difficulty | *Are you deaf, or do you have serious difficulty hearing?*  ● Yes  ● No  ● Prefer not to answer | *Which statement best describes {your/SP's} hearing (without hearing aid)? Would you say {your/his/her} hearing is good, that {you have/s/he has} a little trouble, a lot of trouble, or {are you/is s/he} deaf?*  ● Good (=No in harmonized variable)  ● Little trouble (=No)  ● Lot of trouble (=Yes)  ● Deaf (=Yes)  ● Refused  ● Don't know | ● Yes  ● No |
| Health insurance | *Are you covered by health insurance or some other kind of health care plan?*  ● Yes  ● No  ● Prefer not to answer  ● Don't know | *{Are you/Is SP} covered by health insurance or some other kind of health care plan?*  ● Yes  ● No  ● Refused  ● Don't know | ● Yes  ● No |
| General health | *In general, would you say your health is?*  ● Excellent  ● Very good  ● Good  ● Fair  ● Poor  ● Prefer not to answer | *Would you say {your/SP's} health in general is?*  ● Excellent  ● Very good  ● Good  ● Fair  ● Poor  ● Refused  ● Don't know | ● Excellent  ● Very good  ● Good  ● Fair  ● Poor |
| ***Clinical characteristics*** | | | |
| Asthma | Identified from electronic health records as the presence of a diagnosis prior to the enrollment date.  ● Yes  ● No | *Has a doctor or other health professional ever told {you/SP} that {you have/s/he/SP has} asthma?*  ● Yes  ● No  ● Refused  ● Don't know | ● Yes  ● No |
| Coronary heart disease | Identified from electronic health records as the presence of a diagnosis prior to the enrollment date.  ● Yes  ● No | *Has a doctor or other health professional ever told {you/SP} that {you/s/he} . . .had coronary heart disease?*  ● Yes  ● No  ● Refused  ● Don't know | ● Yes  ● No |
| Congestive heart failure | Identified from electronic health records as the presence of a diagnosis prior to the enrollment date.  ● Yes  ● No | *Has a doctor or other health professional ever told {you/SP} that {you/s/he} . . .had congestive heart failure?*  ● Yes  ● No  ● Refused  ● Don't know | ● Yes  ● No |
| Heart attack | Identified from electronic health records as the presence of a diagnosis prior to the enrollment date.  ● Yes  ● No | *Has a doctor or other health professional ever told {you/SP} that {you/s/he} . . .had a heart attack (also called myocardial infarction)?*  ● Yes  ● No  ● Refused  ● Don't know | ● Yes  ● No |
| Cancer | Identified from electronic health records as the presence of a diagnosis prior to the enrollment date.  ● Yes  ● No | *{Have you/Has SP} ever been told by a doctor or other health professional that {you/s/he} had cancer or a malignancy of any kind?*  ● Yes  ● No  ● Refused  ● Don't know | ● Yes  ● No |
| Hypertension | Identified from electronic health records as the presence of a diagnosis prior to the enrollment date.  ● Yes  ● No | *{Have you/Has SP} ever been told by a doctor or other health professional that {you/s/he} had hypertension, also called high blood pressure?*  ● Yes  ● No  ● Refused  ● Don't know | ● Yes  ● No |
| Obesity | Determined by continuous BMI ≥ 30 kg/m^2^  ● Yes  ● No | Determined by continuous BMI ≥ 30 kg/m^2^  ● Yes  ● No | ● Yes  ● No |
| Stroke | Identified from electronic health records as the presence of a diagnosis prior to the enrollment date.  ● Yes  ● No | *Has a doctor or other health professional ever told {you/SP} that {you/s/he} . . .had a stroke?*  ● Yes  ● No  ● Refused  ● Don't know | ● Yes  ● No |
| Diabetes mellitus | Identified from electronic health records as the presence of a diagnosis prior to the enrollment date.  ● Yes  ● No | *{Other than during pregnancy, {have you/has SP}/{Have you/Has SP}} ever been told by a doctor or health professional that {you have/{he/she/SP} has} diabetes or sugar diabetes?*  ● Yes  ● No  ● Borderline (=missing data in harmonized variable)  ● Refused  ● Don't know | ● Yes  ● No |
| Cataract surgery | Identified from electronic health records as the presence of a procedure code prior to the enrollment date.  ● Yes  ● No | *Have you ever had eye surgery to treat cataracts? (available until 2007-2008)*  ● Yes  ● No  ● Don't know | ● Yes  ● No |

**eAppendix 2. Defining the end of EHR follow-up in All of Us**

In All of Us, the end date of EHR follow-up was determined by the latest administrative censoring date (August 20, 2022) and the probable date of individual censoring. The probable date of individual censoring was based on the most recent date of any electronic health record, plus an observation window. This observation window was set as the 99th percentile of the interval between consecutive clinical visits across all participants, which was 448 days. The assumption was that a participant was no longer at risk of death being observed in EHRs if they had no clinical visit within 448 days after their last recorded visit.

**eAppendix 3. Formulas for calculating population attributable fractions**

We used Levin’s formula to estimate Population Attributable Fractions. Specifically, with a reasonably well-defined reference category, the PAF is defined as

$$PAF= \frac{(\sum_{i=0}^{I} P_{i}\times HR_{i})-1}{\sum_{i=0}^{I} P_{i}\times HR_{i}} ,$$

where $HR_{i}$ is hazard ratio for exposure level $i$ compared with the reference category ($HR_{0}=1$), and $P_{i}$ is the prevalence of exposure level $i$ among the total population. When exposure is binary, the formula reduces to

$$PAF= \frac{P\times(HR-1)}{P\times\left( HR-1 \right)+1} .$$

**eFigure 1. Flow diagram of inclusion criteria in All of Us**

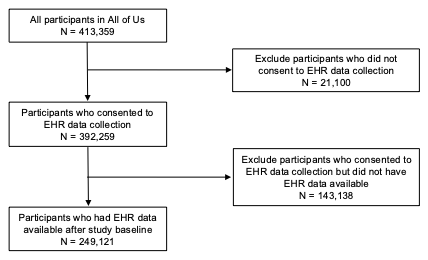

**eFigure 2. Age-specific mortality rates by (A) health and (B) clinical indicators in All of Us**

**A. Mortality rates by general health**

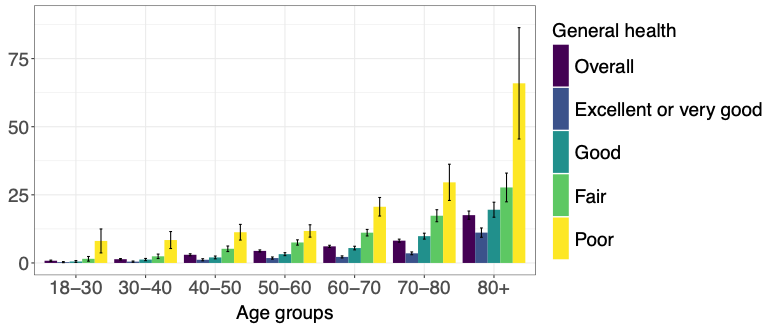

**B. Mortality rates by clinical conditions**

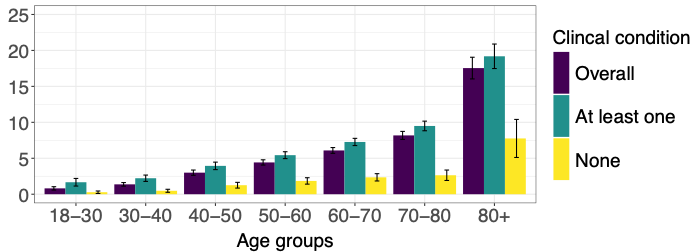

**eFigure 3. Associations of baseline (A) sociodemographic, (B) health-related, and (C) clinical characteristics with mortality in All of Us stratified by levels of healthcare utilization**

**A. Sociodemographic characteristics**

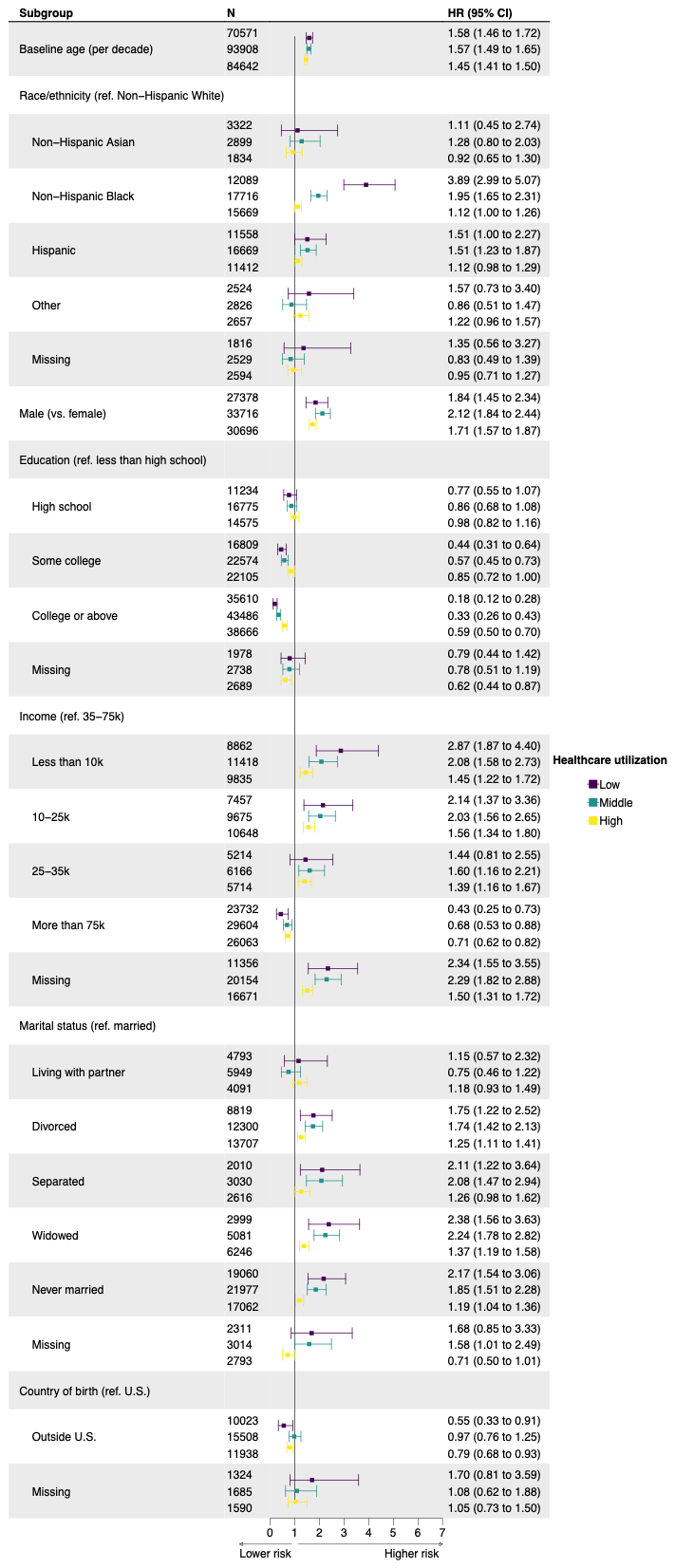

**B. Health-related characteristics**

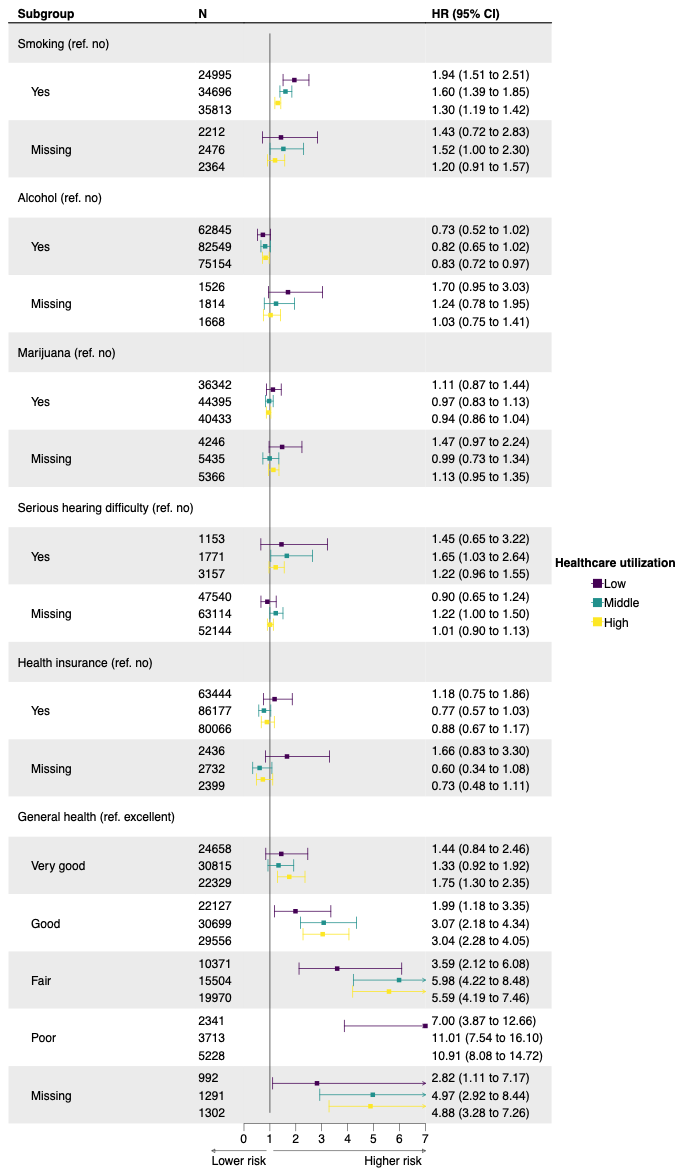

**C. Clinical characteristics**

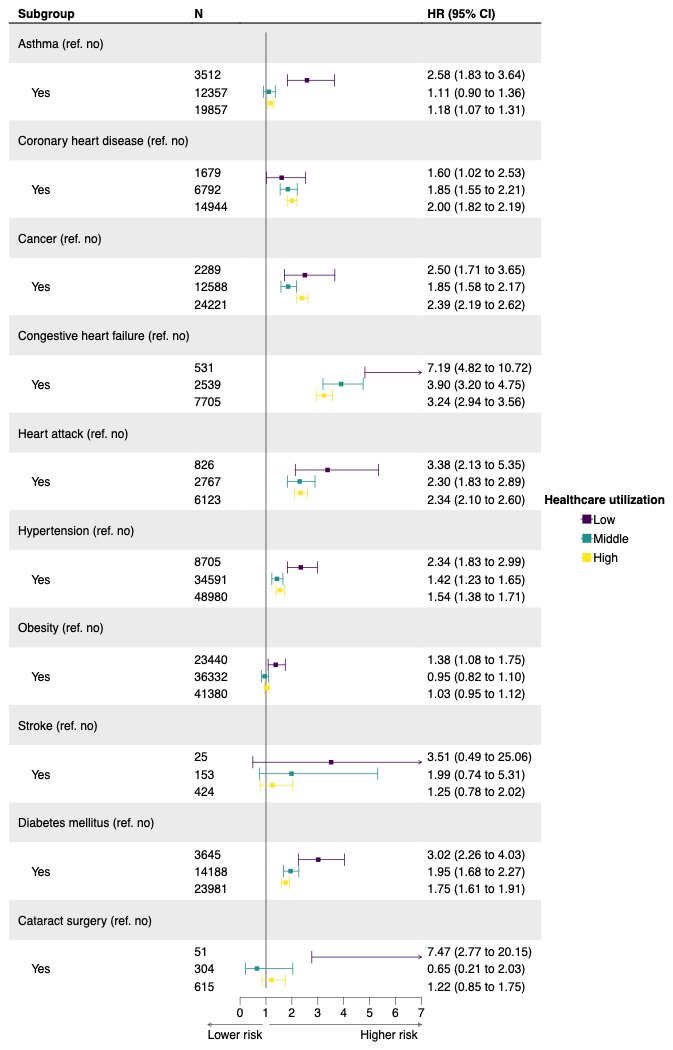

**eTable 1. Diagnostic codes for health-related characteristics in All of Us**

| Characteristics | OMOP concept ID |
| --- | --- |
| Asthma | 317009 |
| Coronary heart disease | 764123, 317576 |
| Congestive heart failure | 4023479, 44782655, 4242669, 4229440, 319835, 314378 |
| Heart attack | 4329847 |
| Cancer | 443392 |
| Hypertension | 312648, 4028741, 320128, 316866, 319826 |
| Stroke | 36684840, 381316, 4153352, 4310996, 4159140 |
| Diabetes mellitus | 201820 |
| Cataract surgery | 4013501, 4223363, 4004519, 4030247, 4208126 |

Abbreviation: OMOP, Observational Medical Outcomes Partnership.

**eTable 2. Baseline characteristics of full All of Us participants**

| Characteristic n (%)^a^ | All of Us  (full sample) |
| --- | --- |
| N | 413359 |
| Mean age (SD) | 51.2 (17.1) |
| Sex |  |
| Female | 249565 (60.4%) |
| Male | 155168 (37.5%) |
| Other or missing | 8626 (2.1%) |
| Race/ethnicity |  |
| Non-Hispanic White | 222645 (53.9%) |
| Non-Hispanic Asian | 13838 (3.3%) |
| Non-Hispanic Black | 77069 (18.6%) |
| Hispanic | 74114 (17.9%) |
| Other | 14001 (3.4%) |
| Missing | 11692 (2.8%) |
| Education |  |
| Less than high school | 36432 (8.8%) |
| High school | 77013 (18.6%) |
| Some college | 104136 (25.2%) |
| College or above | 182345 (44.1%) |
| Missing | 13433 (3.2%) |
| Household income |  |
| <10k | 55544 (13.4%) |
| 10-25k | 47174 (11.4%) |
| 25-35k | 29283 (7.1%) |
| 35-75k | 76128 (18.4%) |
| >75k | 122219 (29.6%) |
| Missing | 83011 (20.1%) |
| Marital status |  |
| Married | 176299 (42.7%) |
| Living with partner | 27712 (6.7%) |
| Divorced | 57008 (13.8%) |
| Separated | 13455 (3.3%) |
| Widowed | 21686 (5.2%) |
| Never married | 102480 (24.8%) |
| Missing | 14719 (3.6%) |
| Country of birth |  |
| U.S. | 343569 (83.1%) |
| Outside U.S. | 62149 (15.0%) |
| Missing | 7641 (1.8%) |
| Smoking history |  |
| Yes | 155900 (37.7%) |
| No | 232630 (56.3%) |
| Missing | 24829 (6.0%) |
| Alcohol history |  |
| Yes | 349996 (84.7%) |
| No | 41630 (10.1%) |
| Missing | 21733 (5.3%) |
| Marijuana history |  |
| Yes | 197797 (47.9%) |
| No | 176912 (42.8%) |
| Missing | 38650 (9.4%) |
| Serious hearing difficulty |  |
| Yes | 10616 (2.6%) |
| No | 149126 (36.1%) |
| Missing | 253617 (61.4%) |
| Health insurance |  |
| Yes | 371633 (89.9%) |
| No | 26778 (6.5%) |
| Missing | 14948 (3.6%) |
| General health |  |
| Excellent | 46352 (11.2%) |
| Very good | 123303 (29.8%) |
| Good | 135334 (32.7%) |
| Fair | 73699 (17.8%) |
| Poor | 17963 (4.3%) |
| Missing | 16708 (4.0%) |
| Asthma |  |
| Yes | n/a |
| No | n/a |
| Missing | n/a |
| Coronary heart disease |  |
| Yes | n/a |
| No | n/a |
| Missing | n/a |
| Congestive heart failure |  |
| Yes | n/a |
| No | n/a |
| Missing | n/a |
| Heart attack |  |
| Yes | n/a |
| No | n/a |
| Missing | n/a |
| Cancer |  |
| Yes | n/a |
| No | n/a |
| Missing | n/a |
| Hypertension |  |
| Yes | n/a |
| No | n/a |
| Missing | n/a |
| Obesity |  |
| Yes | n/a |
| No | n/a |
| Missing | n/a |
| Stroke |  |
| Yes | n/a |
| No | n/a |
| Missing | n/a |
| Diabetes mellitus |  |
| Yes | n/a |
| No | n/a |
| Missing | n/a |
| Cataract surgery |  |
| Yes | n/a |
| No | n/a |
| Missing | n/a |

Abbreviations: SD, standard deviation; n/a, not applicable.

^a^ Percentage may not sum to 100 due to rounding.

**eTable 3. Age-specific mortality rates per 1,000 person years with standard errors by sex in All of Us and continuous NHANES**

| Age groups | Overall | | Female | | Male | |
| --- | --- | --- | --- | --- | --- | --- |
|  | NHANES | All of Us | NHANES | All of Us | NHANES | All of Us |
| 18-30 | 1.1 (0.0) | 0.8 (0.1) | 0.6 (0.0) | 0.6 (0.1) | 1.6 (0.0) | 1.5 (0.3) |
| 30-40 | 1.4 (0.0) | 1.4 (0.1) | 0.8 (0.0) | 1.0 (0.1) | 2.1 (0.0) | 2.1 (0.3) |
| 40-50 | 2.6 (0.0) | 3.0 (0.2) | 2.2 (0.0) | 2.4 (0.2) | 3.0 (0.0) | 4.2 (0.4) |
| 50-60 | 5.5 (0.0) | 4.4 (0.2) | 4.5 (0.0) | 3.3 (0.2) | 6.6 (0.0) | 6.3 (0.4) |
| 60-70 | 11.7 (0.0) | 6.1 (0.2) | 9.2 (0.0) | 4.4 (0.2) | 14.5 (0.0) | 8.7 (0.4) |
| 70-80 | 27.0 (0.0) | 8.2 (0.3) | 21.9 (0.0) | 6.6 (0.3) | 33.4 (0.0) | 10.3 (0.5) |
| 80+ | 93.0 (0.0) | 17.5 (0.8) | 84.8 (0.0) | 13.8 (1.0) | 106.8 (0.0) | 21.8 (1.2) |

**eTable 4. Population attributable fraction for mortality due to characteristics calculated using hazard ratio estimates from All of Us and prevalence estimates from NHANES**

| Characteristic (reference category) | Population attributable fraction |
| --- | --- |
| Education (College or above) | 40.7% |
| Household income (More than 75k) | 39.6% |
| Marital status (Married) | 16.3% |
| Country of birth (Outside U.S.) | 14.4% |
| Smoking history (No) | 17.8% |
| Alcohol history (Yes) | 5.0% |
| Serious hearing difficulty (No) | 1.8% |
| Health insurance (Yes) | 0.1% |
| General health (Excellent) | 68.8% |
| Asthma (No) | 6.6% |
| Coronary heart disease (No) | 4.5% |
| Congestive heart failure (No) | 7.2% |
| Heart attack (No) | 5.9% |
| Cancer (No) | 14.6% |
| Hypertension (No) | 23.1% |
| Obesity (No) | 5.3% |
| Stroke (No) | 2.0% |
| Diabetes mellitus (No) | 9.7% |
| Cataract surgery (No) | 2.4% |
